# Hepatic Toxicity Assessment in HIV’s Interaction with Reverse Transcriptase and Integrase Strand Transfer Inhibitors at a Military Hospital, Southsouth Nigeria

**DOI:** 10.1101/2024.04.30.24306606

**Authors:** Odekunle Bola Odegbemi, Mathew Folaranmi Olaniyan, Musa Abidemi Muhibi

## Abstract

**Introduction:** The use of Anti-retroviral therapy (ART) has made HIV a manageable condition, but there are risks associated with medications like Reverse Transcriptase Inhibitors (RTIs) and Integrase Strand Transfer Inhibitors (INSTIs), such as liver and renal toxicity. It is essential to understand these risks for effective treatment and case management. Investigating liver toxicity related to RTIs and INSTIs in Nigeria is crucial for optimizing HIV treatment. This study aimed to assess the impact of Tenofovir Lamivudine Dolutegravir (TLD) on the liver function of HIV patients at Nigerian Navy Hospital (NNH) Warri.

**Methods:** The liver function of 170 participants was assessed, with 120 on ART and the remainder being HIV-negative attendees at NNH Warri. The study used a cross-sectional design and selected participants through random sampling. We collected data using a semi-structured questionnaire. Blood samples were taken through venipuncture and stored at –20°C before analysis. Ethical approval was obtained, and data analysis was conducted using SPSS Statistical Software Version 23, with significance set at p < 0.05.

**Results:** The study found significant differences in AST, TP, Alb, and GST levels between HIV-positive subjects receiving TLD and HIV-negative individuals. HIV-positive subjects had lower AST and Alb levels but higher TP and GST levels. Further analysis revealed correlations between age, gender, and liver enzymes, highlighting the complex relationship between HIV, liver function, and treatment outcomes.

**Conclusion:** The study suggests that decreased AST levels may have a protective effect, while ALT activity had minimal impact. Changes in TP, Alb, and GSTs emphasize the importance of monitoring hepatic synthetic function and detoxification pathways in HIV patients taking TLD.

## Introduction

Anti-retroviral therapy (ART) has transformed the management of human immunodeficiency virus (HIV), making it a manageable condition. However, medications used in ART, such as Reverse Transcriptase Inhibitors (RTIs) and Integrase Strand Transfer Inhibitors (INSTIs), can pose risks like liver and renal toxicity. The introduction of Tenofovir, Lamivudine, and Dolutegravir (TLD) in Nigeria represents a significant advancement in the country’s efforts to combat HIV/AIDS [1]. This combination of anti-retroviral drugs offers improved treatment outcomes, reduced side effects, and simplified regimens for HIV patients. Its adoption demonstrates Nigeria’s commitment to enhancing access to quality HIV care and aligning with global efforts to control the epidemic.

Tenofovir Lamivudine Dolutegravir (TLD) is a highly effective combination of anti-retroviral drugs that improve treatment outcomes, reduce side effects, and simplify regimens for HIV patients. Its introduction in Nigeria reflects its commitment to improving access to quality HIV care and aligning with global efforts to control the epidemic [2]. Liver toxicity is a significant concern in managing HIV, particularly with the use of ART. Reverse Transcriptase Inhibitors and INSTIs are crucial in suppressing viral replication and improving patient outcomes.

Reverse Transcriptase Inhibitors and INSTIs are vital components of TLD valued for their specificity and manageable toxicity profiles. While first-generation Non-nucleoside RTIs have been associated with liver toxicity, second-generation NNRTIs are considered safer. Understanding these risks is crucial for effective treatment [3]. It is imperative to grasp these nuances for treatment efficacy. Given this, exploring liver toxicity associated with RTIs and INSTIs in Nigeria holds significance for enhancing HIV treatment protocols.

Liver toxicity poses a significant concern in HIV management, particularly with ART usage. RTIs and INSTIs are crucial in suppressing viral replication and improving patient outcomes. However, emerging evidence suggests that these drugs may have hepatotoxic effects, which complicate their clinical use. In Nigeria, where HIV/AIDS is a priority, understanding the prevalence and patterns of liver toxicity related to RTIs and INSTIs is crucial for treatment optimization and minimizing adverse outcomes in HIV-infected individuals [4; 5].

This research aims to clarify the dynamics of liver injury associated with TLD, providing valuable information for clinical decisions, patient monitoring, and improving HIV management in resource-limited settings. The study specifically aims to explore the incidence, severity, and risk factors associated with liver toxicity among HIV patients receiving TLD at a military hospital in Southsouth Nigeria. Like other government healthcare facilities, military hospitals serve a diverse patient population, including active-duty personnel, veterans, and civilians. They provide a unique setting to investigate the intersection of HIV infection and liver toxicity.

## Methods

In this study, a cross-sectional design was utilized, and a carefully selected group of participants was included. The group consisted of 120 individuals who were HIV-positive and receiving TLD. Another 50 individuals who were HIV-negative and receiving care at Nigerian Navy Hospital (NNH) Warri were recruited as controls. We matched the controls with the HIV-positive group in terms of sex, age, socioeconomic status, and ethnicity to minimize potential influences on the study results and ensure the validity of the findings. NNH Warri, located in Uvwie Local Government Area of Delta State, primarily serves Nigerian military and paramilitary personnel and their families. Additionally, NNH Warri extends its healthcare services to the general public as part of its social responsibility initiatives. It is also a Ministry of Defence Health Implementation Program site for the United States Department of Defense Program on HIV/AIDS in Delta State, Nigeria.

The study enrolled consenting adult male and female Nigerians between the ages of 18 and 65. Individuals with primary chronic conditions such as liver cirrhosis, chronic obstructive pulmonary disease, cancer, and alcoholism were excluded from the study. We employed simple random sampling to ensure the representation of diverse demographic elements in the study. Ethical approval was obtained from the Ministry of Defence Research and Ethics Review Committee (NHREC/MOD-HREC/15/02/23C), and administrative approval was obtained from the management of NNH Warri prior to the commencement of the study. Data collection involved administering a semi-structured questionnaire to the participants, allowing for a detailed exploration of their clinical, demographic, and behavioral characteristics.

Aseptic venipuncture was used to collect five milliliters (5ml) of whole blood samples, which were then placed in a Lithium Heparin specimen bottle. Medical laboratory professionals utilized sterile disposable vacutainer needles for collecting the blood samples. The samples were subsequently centrifuged at 4,000 revolutions per minute for 3-5 minutes to obtain plasma, which was stored at –20°C until laboratory analysis. We used relevant biochemical methods to determine the plasma levels of liver function parameters, including Alanine Aminotransferase (ALT), Aspartate Aminotransferase (AST), Glutathione S-transferases (GSTs), Total Protein (TP), and Albumin (Alb). Control samples and commercial standards provided by kit manufacturers were tested alongside each batch of plasma samples to ensure the accuracy of the test kits. Prior to analysis, the collected data underwent a cleaning process.

Data obtained were entered into SPSS statistical software version 23, and a significance level of p < 0.05 was utilized. This comprehensive approach, encompassing participant selection, data collection, and analysis, enhances the reliability and relevance of the study’s findings, offering valuable insights into the dynamics of HIV within the study population.

## Results

Findings from the study are presented in this section. Table 1 displays the socio-demographic characteristics of all study participants, aged 18 to 65 years, with a mean age of 38.6 ± 11.2 years. Among the 170 participants, 94 (55.3%) were female, of whom 68 (40.0%) were HIV positive. The majority were married (47.0%), from monogamous families (63.5%), and of Urhobo/Itsekiri ethnicity (45.2%). Educationally, 91 (53.5%) had secondary education, 41 (24.1%) had tertiary education, and 21 (12.3%) had primary education. Trading was the predominant occupation (50.6%), followed by civil service (16.5%).

**Table 1:** Demographic Characteristics of Study Participants.

| Variables | Frequency (n = 170) | Percentage (%) |
| --- | --- | --- |
| **Age Group (years)** |  |  |
| ≤20 | 11 | 6.4 |
| 21-30 | 33 | 19.4 |
| 31-40 | 50 | 29.4 |
| 41-50 | 60 | 35.5 |
| 51-60 | 8 | 4.7 |
| >60 | 8 | 4.7 |
| Mean ± SD | 38.6 ± 11.2 |  |
| **Gender** |  |  |
| Male | 76 | 44.7 |
| Female | 94 | 55.3 |
| **Marital Status** |  |  |
| Single | 62 | 36.4 |
| Married | 80 | 47.0 |
| Co-habiting | 11 | 6.5 |
| Widowed | 2 | 1.2 |
| Divorced/separated | 8 | 4.7 |
| Missing | 7 | 4.1 |
| **Type of family** |  |  |
| Monogamy | 108 | 63.5 |
| Polygamy | 57 | 33.5 |
| Missing | 5 | 2.9 |
| **Ethnic Group** |  |  |
| Hausa/Fulani | 22 | 12.9 |
| Yoruba | 28 | 22.3 |
| Igbo | 37 | 21.8 |
| Others | 77 | 45.2 |
| Missing | 6 | 3.5 |
| **Religion** |  |  |
| Christianity | 127 | 74.7 |
| Islam | 32 | 18.8 |
| Traditional | 8 | 4.7 |
| Missing | 3 | 1.8 |
| **Educational Level** |  |  |
| Primary | 21 | 12.3 |
| Secondary | 91 | 53.5 |
| Tertiary | 41 | 24.1 |
| Postgraduate | 4 | 2.4 |
| None | 2 | 1.2 |
| Missing | 11 | 6.4 |
| **Occupation** |  |  |
| Student | 25 | 14.7 |
| Civil servant | 28 | 16.5 |
| Farmer | 2 | 1.2 |
| Business/Trader | 86 | 50.6 |
| Unemployed | 8 | 4.7 |
| Retired | 8 | 4.7 |
| Missing | 5 | 2.9 |

Table 2 outlines the characteristics of HIV seropositive individuals, with an average age of 38.4 ± 11.1 years. Among the 120 participants, 68 (56.7%) were female, and 57 (47.5%) were married. Predominantly from the Urhobo/Itsekiri ethnic group (50.8%), most had secondary education (55.8%) and were engaged in trading (50.8%). Table 3 presents data on HIV seronegative individuals with an average age of 39.0 ± 11.3 years. Among the 50 participants, 26 (52.0%) were female, and 23 (46.0%) were married. The majority were from monogamous families (58.0%) and practiced Christianity (62.0%). The Urhobo/Itsekiri ethnic group was also prominent (36.0%), with most having secondary education (46.0%) and engaged in trading (50.0%).

**Table 2:**
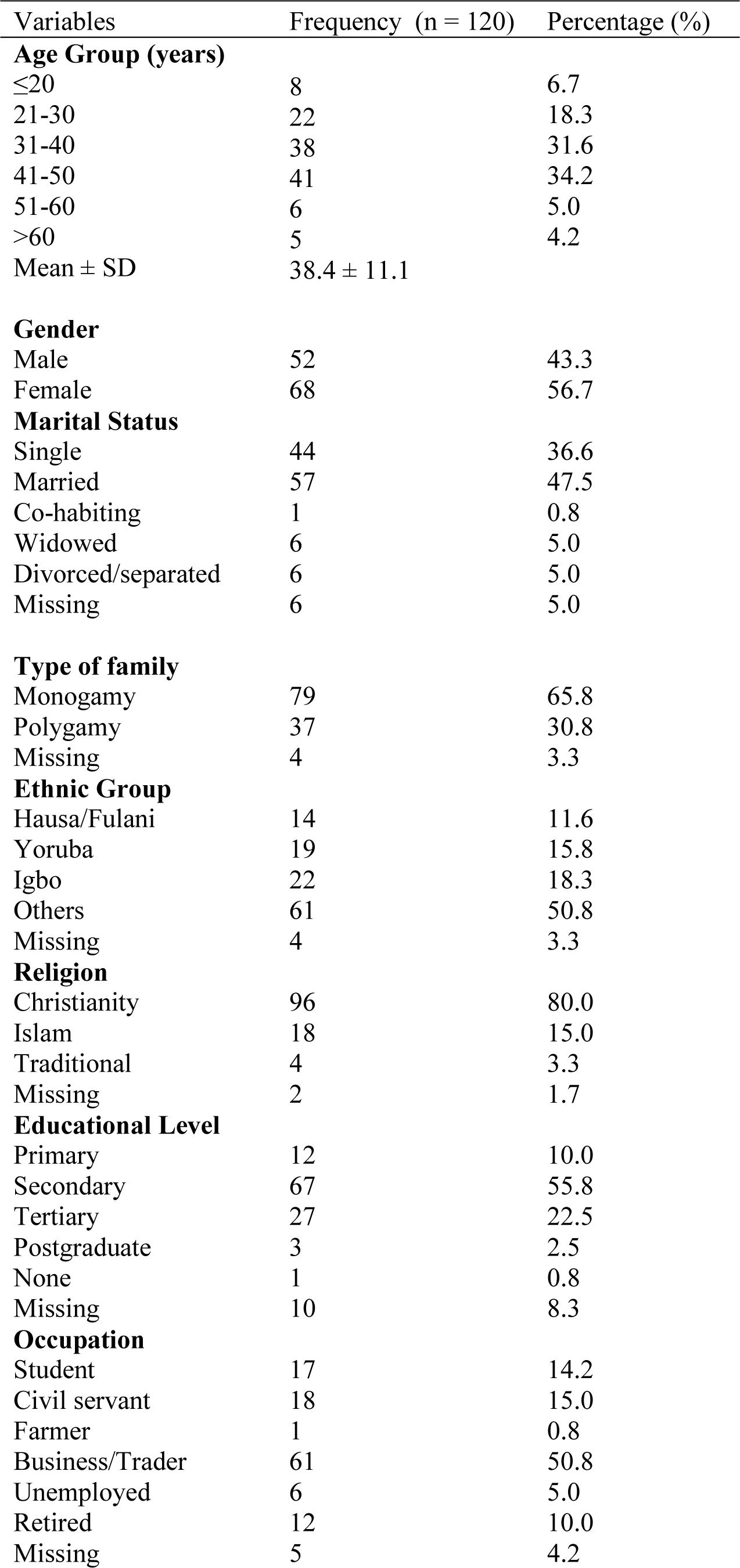
Demographic Characteristics of HIV Seropositive Subjects at Nigerian Navy Hospital Warri.

| Variables | Frequency (n = 120) | Percentage (%) |
| --- | --- | --- |
| **Age Group (years)** |  |  |
| ≤20 | 8 | 6.7 |
| 21-30 | 22 | 18.3 |
| 31-40 | 38 | 31.6 |
| 41-50 | 41 | 34.2 |
| 51-60 | 6 | 5.0 |
| >60 | 5 | 4.2 |
| Mean ± SD | 38.4 ± 11.1 |  |
| **Gender** |  |  |
| Male | 52 | 43.3 |
| Female | 68 | 56.7 |
| **Marital Status** |  |  |
| Single | 44 | 36.6 |
| Married | 57 | 47.5 |
| Co-habiting | 1 | 0.8 |
| Widowed | 6 | 5.0 |
| Divorced/separated | 6 | 5.0 |
| Missing | 6 | 5.0 |
| **Type of family** |  |  |
| Monogamy | 79 | 65.8 |
| Polygamy | 37 | 30.8 |
| Missing | 4 | 3.3 |
| **Ethnic Group** |  |  |
| Hausa/Fulani | 14 | 11.6 |
| Yoruba | 19 | 15.8 |
| Igbo | 22 | 18.3 |
| Others | 61 | 50.8 |
| Missing | 4 | 3.3 |
| **Religion** |  |  |
| Christianity | 96 | 80.0 |
| Islam | 18 | 15.0 |
| Traditional | 4 | 3.3 |
| Missing | 2 | 1.7 |
| **Educational Level** |  |  |
| Primary | 12 | 10.0 |
| Secondary | 67 | 55.8 |
| Tertiary | 27 | 22.5 |
| Postgraduate | 3 | 2.5 |
| None | 1 | 0.8 |
| Missing | 10 | 8.3 |
| **Occupation** |  |  |
| Student | 17 | 14.2 |
| Civil servant | 18 | 15.0 |
| Farmer | 1 | 0.8 |
| Business/Trader | 61 | 50.8 |
| Unemployed | 6 | 5.0 |
| Retired | 12 | 10.0 |
| Missing | 5 | 4.2 |

**Table 3:** Demographic Characteristics of HIV Seronegative Subjects Attending Nigerian Navy Hospital Warri.

| Variables | Frequency (n = 50) | Percentage (%) |
| --- | --- | --- |
| **Age Group (years)** |  |  |
| ≤20 | 3 | 6.0 |
| 21-30 | 11 | 22.0 |
| 31-40 | 12 | 24.0 |
| 41-50 | 19 | 38.0 |
| 51-60 | 2 | 4.0 |
| >60 | 3 | 6.0 |
| Mean ± SD | 39.0 ± 11.3 |  |
| **Gender** |  |  |
| Male | 24 | 48.0 |
| Female | 26 | 52.0 |
| **Marital Status** |  |  |
| Single | 18 | 36.0 |
| Married | 23 | 46.0 |
| Co-habiting | 1 | 2.0 |
| Widowed | 5 | 10.0 |
| Divorced/separated | 2 | 4.0 |
| Missing | 1 | 2.0 |
| **Type of family** |  |  |
| Monogamy | 29 | 58.0 |
| Polygamy | 20 | 40.0 |
| Missing | 1 | 2.0 |
| **Ethnic Group** |  |  |
| Hausa/Fulani | 8 | 16.0 |
| Yoruba | 9 | 18.0 |
| Igbo | 15 | 30.0 |
| Others | 18 | 36.0 |
| Missing | 2 | 4.0 |
| **Religion** |  |  |
| Christianity | 31 | 62.0 |
| Islam | 14 | 28.0 |
| Traditional | 4 | 8.0 |
| Missing | 1 | 2.0 |
| **Educational Level** |  |  |
| Primary | 10 | 20.0 |
| Secondary | 23 | 46.0 |
| Tertiary | 14 | 28.0 |
| Postgraduate | 1 | 2.0 |
| None | 1 | 2.0 |
| Missing | 1 | 2.0 |
| **Occupation** |  |  |
| Student | 8 | 16.0 |
| Civil servant | 10 | 20.0 |
| Farmer | 1 | 2.0 |
| Business/Trader | 25 | 50.0 |
| Unemployed | 2 | 4.0 |
| Retired | 1 | 2.0 |
| Missing | 3 | 6.0 |

Table 4 shows independent sample t-tests, comparing the mean levels of various liver function markers between the two groups: HIV Seropositive Subjects on TLD and HIV Seronegative Subjects attending NNH Warri. A notable statistical variance was detected in the mean levels of AST, TP, Alb, and GSTs among the subjects. The mean values of AST and Alb were lower in HIV subjects when compared with HIV seronegative subjects. In comparison, the values of TP and GSTs were significantly higher in HIV subjects when compared to HIV seronegative subjects.

**Table 4:** Effect of TLD on Liver Function Parameters Among HIV Seropositive at NNH Warri.

|  | Group | N | Mean | Std. Dev | SEM | F | Ta | Df | p-value |
| --- | --- | --- | --- | --- | --- | --- | --- | --- | --- |
| AST (IU/l) | HIV + | 120 | 21.78 | 7.300 | 0.666 | 1.829 | -2.350 | 75.51 | .010* |
|  | HIV - | 50 | 25.24 | 9.270 | 1.311 |  |  |  |  |
| ALT (IU/l) | HIV + | 120 | 30.73 | 13.15 | 1.201 | 4.095 | .655 | 124.8 | .256 |
|  | HIV - | 50 | 29.54 | 9.558 | 1.352 |  |  |  |  |
| TP (g/dl) | HIV + | 120 | 8.674 | 0.813 | 0.074 | 2.018 | 7.744 | 116.1 | .000* |
|  | HIV - | 50 | 7.770 | 0.637 | 0.090 |  |  |  |  |
| Alb (g/dl) | HIV + | 120 | 3.221 | 0.985 | 0.090 | 12.30 | -12.44 | 142.2 | .000* |
|  | HIV - | 50 | 4.782 | 0.618 | 0.087 |  |  |  |  |
| GSTs (ng/ml) | HIV + | 120 | 5.106 | 2.617 | 0.239 | 6.253 | 2.998 | 150.7 | .001* |
|  | HIV - | 50 | 4.144 | 1.514 | 0.214 |  |  |  |  |

<sup>a</sup> Equal variance not assumed in the t-test
\* p-value is statistically significant at 95% confidence interval

Table 5 shows correlation coefficients between the various variables in the study, along with their corresponding p-values, indicating the significance of these correlations. Age showed a negative, weak correlation with Gender (–0.063, p = 0.494). The duration of ART intake had a negative, moderate correlation with the age at which ART started (–0.184, p = 0.047). This suggests that older participants tend to have started ART later. AST and ALT showed positive correlations (0.241, p = 0.009). AST and ALT negatively correlated with total protein (TP) and albumin (Alb). AST and ALT also had positive correlations with some other variables. Age positively correlated with AST (0.137, p = 0.135). This indicates a slight tendency for older individuals to have higher levels of AST. Age negatively correlated with ALT (–0.066, p = 0.474). This suggests a very slight tendency for older individuals to have lower levels of ALT. Gender does not significantly correlate with AST (–0.131, p = 0.153). Based on this data, no definite correlation was observed between gender and AST levels. Gender significantly correlated with ALT (0.000, p = 0.997). The weak correlation suggested no clear association between gender and ALT levels.

**Table 5:** Pearson Correlations Between Sociodemographic Characteristics Years of Therapy, and Liver Function Variables Among HIV Seropositive Subjects Attending NNH Warri.

|  |  | Age | Gender | When ART Started | AST (IU/l) | ALT (IU/l) | TP (g/dl) | Alb (g/dl) | GSTs (ng/ml) |
| --- | --- | --- | --- | --- | --- | --- | --- | --- | --- |
| **Age** | **Correlation** | 1 | -.063 | -.184* | .137 | -.066 | -.067 | .013 | .100 |
|  | **p-value** |  | .494 | .047 | .135 | .474 | .468 | .888 | .279 |
|  | **N** | 120 | 120 | 118 | 120 | 120 | 120 | 120 | 120 |
| **Gender** | **Correlation** | -.063 | 1 | -.038 | -.131 | .000 | .007 | .009 | -.037 |
|  | **p-value** | .494 |  | .679 | .153 | .997 | .938 | .922 | .685 |
|  | **N** | 120 | 120 | 118 | 120 | 120 | 120 | 120 | 120 |
| **When ART started** | **Correlation** | -.184* | -.038 | 1 | -.088 | .205* | .084 | .147 | -.010 |
|  | **p-value** | .047 | .679 |  | .344 | .026 | .366 | .113 | .918 |
|  | **N** | 118 | 118 | 118 | 118 | 118 | 118 | 118 | 118 |
| **AST (IU/l)** | **Correlation** | .137 | -.131 | -.088 | 1 | -.258* | -.247* | .177 | -.169 |
|  | **P-value** | .135 | .153 | .344 |  | .004 | .007 | .056 | .066 |
|  | **N** | 120 | 120 | 118 | 120 | 120 | 120 | 118 | 120 |
| **ALT (IU/l)** | **Correlation** | -.066 | .000 | .205* | -.258* | 1 | .272** | -.187* | .132 |
|  | **P-value** | .474 | .997 | .026 | .004 |  | .003 | .040 | .149 |
|  | **N** | 120 | 120 | 118 | 120 | 120 | 120 | 120 | 120 |
| **TP (g/dl)** | **Correlation** | -.067 | .007 | .084 | -.247* | .272* | 1 | .866* | -.087 |
|  | **P-value** | .468 | .938 | .366 | .007 | .003 |  | .000 | .347 |
|  | **N** | 120 | 120 | 118 | 120 | 120 | 120 | 120 | 120 |
| **Alb (g/dl)** | **Correlation** | .100 | -.037 | -.010 | -.169 | .132 | -.087 | .228* | 1 |
|  | **P-value** | .279 | .685 | .918 | .066 | .149 | .347 | .012 |  |
|  | **N** | 120 | 120 | 118 | 120 | 120 | 120 | 120 | 120 |
| **GSTs (ng/ml)** | **Correlation** | .013 | .009 | .147 | -.187* | .866** | .228* | .540 | .057 |
|  | **P-value** | .888 | .922 | .113 | .040 | .000 | .012 | 120 | .540 |
|  | **N** | 120 | 120 | 118 | 120 | 120 | 120 | 1 | 120 |

\*. Correlation is significant at the 0.05 level (2-tailed).

Table 6 presents the relationship between different liver function parameters among individuals who were HIV-negative. We observed a moderate positive correlation (0.362) between AST and ALT, indicating that both enzymes tend to increase simultaneously. This correlation is expected since both enzymes elevate in response to liver damage. A strong positive correlation (0.634) between ALT and GSTs suggests that higher ALT levels are often associated with higher GST levels. Additionally, a strong positive correlation (0.654) was found between TP and Alb, indicating that higher levels of total protein generally coincide with higher levels of albumin. However, no statistically significant correlations were found between AST and Alb, TP and ALT, TP and GSTs, or Alb and GSTs.

**Table 6:** Pearson Correlations Between Liver Function Variables Among HIV Seronegative Subjects Attending NNH Warri.

|  |  | AST (IU/l) | ALT (IU/l) | TP (g/dl) | Alb (g/dl) | GSTs (ng/ml) |
| --- | --- | --- | --- | --- | --- | --- |
| AST (IU/l) | Correlation | 1 | 0.362* | -0.079 | -0.058 | 0.084 |
|  | Sig. (2-tailed) |  | 0.010 | 0.584 | 0.691 | 0.561 |
|  | N | 50 | 50 | 50 | 50 | 50 |
| ALT (IU/l) | Correlation | 0.362* | 1 | -0.166 | -0.042 | 0.634* |
|  | Sig. (2-tailed) | 0.010 |  | 0.250 | 0.771 | 0.000 |
|  | N | 50 | 50 | 50 | 50 | 50 |
| TP (g/dl) | Correlation | -0.079 | -0.166 | 1 | 0.654* | 0.098 |
|  | Sig. (2-tailed) | 0.584 | 0.250 |  | 0.000 | 0.499 |
|  | N | 50 | 50 | 50 | 50 | 50 |
| Alb (g/dl) | Correlation | -0.058 | -0.042 | 0.654* | 1 | 0.130 |
|  | Sig. (2-tailed) | 0.691 | 0.771 | 0.000 |  | 0.370 |
|  | N | 50 | 50 | 50 | 50 | 50 |
| GSTs (ng/ml) | Correlation | 0.084 | 0.634* | 0.098 | 0.130 | 1 |
|  | Sig. (2-tailed) | 0.561 | 0.000 | 0.499 | 0.370 |  |
|  | N | 50 | 50 | 50 | 50 | 50 |

\*. Correlation is significant at the 0.05 level (2-tailed).

## Discussion

Findings from this study showed a statistically significant difference in the mean levels of AST, TP, Alb, and GSTs among the subjects. The statistical significance observed in the mean levels of these liver function markers between the two groups suggests that TLD treatment may have a notable impact on liver function parameters in HIV seropositive individuals [6; 7]. Understanding these differences is crucial for healthcare providers managing HIV patients, as liver function plays a vital role in overall health and treatment outcomes.

Aspartate Aminotransferase levels showed a considerable difference between individuals with HIV seropositive on TLD and HIV seronegative subjects, with the former group showing lower levels. This decrease in AST levels among individuals receiving TLD suggests a potential protective effect of this anti-retroviral therapy on hepatic integrity [8]. However, further investigations are necessary to elucidate the mechanisms underlying this observation and its clinical implications.

In contrast, no significant difference was observed in ALT levels between HIV seropositive individuals on TLD and HIV seronegative subjects. Alanine Aminotransferase is a crucial enzyme indicating liver injury, and the comparable levels between the two groups suggest that TLD treatment may not significantly affect ALT activity in HIV-positive individuals [9]. Nevertheless, continuous monitoring of ALT levels is essential to detect potential hepatotoxic effects associated with TLD therapy.

The significant difference in TP levels suggests alterations in hepatic protein synthesis among HIV-positive individuals on TLD [10]. Elevated TP levels imply modifications in hepatic protein metabolism secondary to anti-retroviral therapy, necessitating further exploration of specific proteins involved and their clinical implications.

Similarly, the significant difference in plasma Alb levels highlights the impact of TLD treatment on hepatic albumin synthesis. Lower Alb levels in HIV-positive subjects on TLD suggest potential hepatocellular dysfunction or impaired albumin production related to anti-retroviral therapy [11]. These findings, similar to those reported by Abba et al. in Cameroon, underscore the importance of comprehensive assessment and monitoring of hepatic synthetic function in HIV patients on TLD [12].

Moreover, the significant difference in GST levels suggests alterations in hepatic detoxification pathways among HIV-positive individuals [13]. Elevated GST levels, particularly in those on TLD, indicate increased hepatic oxidative stress and cellular injury, emphasizing the need to evaluate hepatic antioxidant capacity and detoxification mechanisms in HIV patients on TLD.

Outcomes of independent sample t-tests comparing liver function markers between the groups, revealed significant differences in AST, TP, Alb, and GST levels as shown in Table 4. The lower mean values of AST and Alb in HIV-positive subjects suggest potential liver dysfunction compared to HIV-negative individuals. Conversely, higher mean values of TP and GSTs in HIV-positive subjects indicate possible liver stress or inflammation due to HIV infection.

These findings align with previous studies that have reported altered liver function in HIV-positive individuals. For instance, a study by Dusingize et al. found that HIV-positive patients had significantly higher levels of liver enzymes, such as AST and ALT, compared to HIV-negative controls, suggesting liver dysfunction in HIV-positive individuals [14]. However, a study by Osakunor et al. in Ghana reported no significant change in liver enzymes among ART clients, with a caveat that liver enzyme monitoring should be ensured among HIV clients despite the findings [15].

Age exhibited a modest association with liver enzymes, while gender showed minimal correlation, emphasizing the need for further research and clinical context to understand these relationships fully. The strong positive correlation between ALT and GSTs suggests a robust association between liver enzyme elevation and oxidative stress, warranting additional investigation into potential liver damage.

The correlation analysis in Table 5 provides insights into the relationships between variables, aiding in contextualizing the results and identifying consistencies across different populations. The findings contribute to our understanding of liver function in HIV/AIDS patients, guiding clinical management and future research.

The negative and weak correlation observed between age and gender in this study is consistent with prior findings [16; 17], indicating that the relationship between age and gender may not be substantial within HIV/AIDS cohorts. In the same vein, the negative, moderate correlation between the duration of ART intake and the age of starting ART is consistent with findings from other studies [18; 19], indicating that older individuals may initiate ART later in their disease course.

The positive correlation observed between AST and ALT levels is a common finding in studies assessing liver function in HIV/AIDS patients [20; 21; 22]. This suggests that elevations in one liver enzyme may be accompanied by elevations in the other, reflecting potential liver injury or inflammation. However, Wood et al. 2021 reported that long-term use of INSTI-based ART reduces liver enzyme elevation by half compared to those on NNRTI [23]. This observed difference may, however, be due to differences in race, ethnicity, and study design.

The weak positive correlation between age and AST (0.137) suggests a slight tendency for older patients to have higher AST levels. This finding is not conclusive and requires further investigation with a larger sample size. Interestingly, a weak negative correlation between age and ALT (–0.066) was observed, contradicting what was reported by [24]. No significant correlations were found between gender and either AST or ALT levels. This aligns with some studies suggesting minimal influence of gender on these enzymes in healthy individuals [25].

Furthermore, the negative correlations between AST, ALT, and other markers of liver function, such as TP and Alb, are consistent with existing literature [26; 27], highlighting the impact of liver dysfunction on these biochemical parameters. Regarding the correlations involving age and gender with AST and ALT levels, the findings may vary across studies. While some studies may report similar trends [14], others may find conflicting results [28], emphasizing the need for further investigation into the influence of age and gender on liver enzyme levels in HIV/AIDS patients.

Accordingly, the correlations identified in this study contribute to our understanding of liver function in HIV/AIDS patients receiving ART. We can discern patterns and discrepancies by comparing these results with similar studies, ultimately guiding clinical management and future research endeavors. The moderate positive correlation between AST and ALT in Table 6 aligns with established knowledge, suggesting similar mechanisms for liver enzyme elevation in HIV-positive individuals [20].

The strong positive correlation (0.634) observed between ALT and GSTs among HIV seronegative subjects is an exciting finding. While not directly explored in the context of HIV, increased GST levels can be a compensatory response to oxidative stress, which can occur in liver damage [29]. Further research is needed to understand this association better. The strong positive correlation (0.654) between TP and Alb is consistent with expectations, as albumin is the major protein produced by the liver [30].

Both groups showed positive correlations between AST and ALT, suggesting similar underlying mechanisms for liver enzyme elevation. However, the study design cannot definitively determine if HIV or ART medication contributes to the observed correlations in the HIV-positive group. This study underscores the importance of monitoring liver function in HIV-positive individuals and addressing potential complications associated with HIV and anti-retroviral therapy. Continued research is essential to elucidate the mechanisms underlying liver dysfunction and develop targeted interventions to improve liver health in this population.

## Conclusion

Our study reveals significant differences in AST, TP, Alb, and GST levels among HIV-positive individuals, suggesting TLD treatment’s impact on liver function. While TLD seems to lower AST levels, implying liver protection [31], further research is needed to understand its mechanisms. Conversely, ALT levels showed no significant change, indicating minimal impact on liver activity. Continuous monitoring remains crucial to detect potential hepatotoxic effects of TLD therapy.

The notable differences in TP and Alb levels underscore alterations in hepatic protein synthesis among HIV seropositive individuals on TLD [32]. Elevated TP levels suggest modifications in hepatic protein metabolism secondary to anti-retroviral therapy, while lower Alb levels may indicate hepatocellular dysfunction [10]. Similar findings reported in Cameroon emphasize the importance of comprehensive assessment and monitoring of hepatic synthetic function in HIV patients on TLD [12]. Moreover, significant differences in GST levels highlight alterations in hepatic detoxification pathways [33], necessitating the evaluation of antioxidant capacity and detoxification mechanisms in HIV patients receiving TLD.

Comparing our findings with previous research suggests consistency in associations between age, gender, and liver enzymes [16; 17]. The observed positive correlation between AST and ALT levels aligns with existing literature on liver function in HIV/AIDS patients [21; 22], indicating potential liver injury or inflammation. However, differences in enzyme elevation between INSTI-based and NNRTI-based ART regimens warrant further investigation [34]. The negative correlations between AST, ALT, and other liver markers, along with the varying associations of age and gender, contribute to our understanding of liver function in this population [26; 27].

Reverse Transcriptase Inhibitors and INSTI therapy benefits HIV clients, but metabolic syndrome and associated liver disease pose growing risks as HIV-positive individuals age [34]. The correlations in this study offer insights into the intricate relationship between HIV, ART, and liver function, guiding clinical care and future research. However, our study had limitations, including a small sample size and a lack of exploration of potential confounding factors like other viral infections among paticipants. These may have had impact on our findings Longitudinal studies are needed to uncover mechanisms behind liver dysfunction in HIV patients and develop targeted interventions for hepatic health improvement.

## Data Availability

All data obtained in the present study are available upon reasonable request to the corresponding author.

## Acknowledgment

The Nigerian Ministry of Defence Health Implementation Programme and the Management of Nigerian Navy Hospital Warri are appreciated for providing the enabling environment and approving this study.

## Author Contributions

OBO participated in research design, research performance, data analysis, and writing of the paper.

MFO participated in research design, research supervision, data analysis, and writing of the paper.

MAA participated in the research design and performance of research.

